# Personalized Tacrolimus Dosing After Liver Transplantation: A Randomized Clinical Trial

**DOI:** 10.1101/2023.05.26.23290604

**Authors:** Jeffrey Khong, Megan Lee, Curtis Warren, Un Bi Kim, Sergio Duarte, Kenneth A. Andreoni, Sunaina Shrestha, Mark W. Johnson, Narendra R. Battula, Danielle M. McKimmy, Thiago Beduschi, Ji-Hyun Lee, Derek M. Li, Chih-Ming Ho, Ali Zarrinpar

## Abstract

**Background:** Inter- and intra-individual variability in tacrolimus dose requirements mandates empirical clinician-titrated dosing that frequently results in deviation from a narrow target range. Improved methods to individually dose tacrolimus are needed. Our objective was to determine whether a quantitative, dynamically-customized, phenotypic-outcome-guided dosing method termed Phenotypic Personalized Medicine (PPM) would improve target drug trough maintenance.

**Methods:** In a single-center, randomized, pragmatic clinical trial (NCT03527238), 62 adults were screened, enrolled, and randomized prior to liver transplantation 1:1 to standard-of-care (SOC) clinician-determined or PPM-guided dosing of tacrolimus. The primary outcome measure was percent days with large (>2 ng/mL) deviation from target range from transplant to discharge. Secondary outcomes included percent days outside-of-target-range and mean area-under-the-curve (AUC) outside-of-target-range per day. Safety measures included rejection, graft failure, death, infection, nephrotoxicity, or neurotoxicity.

**Results:** 56 (29 SOC, 27 PPM) patients completed the study. The primary outcome measure was found to be significantly different between the two groups. Patients in the SOC group had a mean of 38.4% of post-transplant days with large deviations from target range; the PPM group had 24.3% of post-transplant days with large deviations; (difference -14.1%, 95% CI: -26.7 to -1.5 %, P=0.029). No significant differences were found in the secondary outcomes. In post-hoc analysis, the SOC group had a 50% longer median length-of-stay than the PPM group [15 days (Q1-Q3: 11-20) versus 10 days (Q1-Q3: 8.5-12); difference 5 days, 95% CI: 2-8 days, P=0.0026].

**Conclusions:** PPM guided tacrolimus dosing leads to better drug level maintenance than SOC. The PPM approach leads to actionable dosing recommendations on a day-to-day basis.

**Lay Summary:** In a study on 62 adults who underwent liver transplantation, researchers investigated whether a new dosing method called Phenotypic Personalized Medicine (PPM) would improve daily dosing of the immunosuppression drug tacrolimus. They found that PPM guided tacrolimus dosing leads to better drug level maintenance than the standard-of-care clinician-determined dosing. This means that the PPM approach leads to actionable dosing recommendations on a day-to-day basis and can help improve patient outcomes.

## Introduction

Tacrolimus is the most commonly used immunosuppression drug after solid organ transplantation.[1-3] However, its narrow therapeutic window[4] and high inter- and intra-individual variability in dosing requirements, particularly across diverse patient populations, necessitates clinician-titrated dosing that often results in deviation from target ranges, particularly during the critical post-operative phase.[5-7] High intra-patient variability in blood tacrolimus levels may be associated with poor outcomes, including rejection and graft loss.[8-12] Appropriate dosing of tacrolimus during the first 2 weeks after liver transplantation have further been associated with lower risk of graft loss.[13] Therefore, there is a clear need for personalized medicine to address post-transplant immunosuppression by improving therapeutic consistency and augmenting clinical decision making. Until recently, a robust procedure to achieve personalized dosing of tacrolimus and other post-operative drugs has not been available.[14]

Post-transplant immunosuppression provides a challenging model to test any personalized medicine platform. Previous attempts at tacrolimus dosing have used genetics, pharmacokinetics, and other predictive models.[15-18] However, it has proven difficult to simultaneously account for inter- and intra-individual variability in treatment regimens using such approaches, much less to use them to dose patients on a day-to-day basis. These differences also lead to health disparities not solely attributable to access, economics, or adherence.[19-21] Tacrolimus is a substrate of cytochrome P450 and P-glycoprotein, proteins with variable activity in intestine and liver.[6, 22, 23] Its clearance is dependent on liver and kidney function, which can vary tremendously in the post-transplant setting.[24] Furthermore, transplant patients take multiple interacting medications. Current methods based on population-averaged pharmacodynamics, pharmacogenetics, or pharmacokinetics cannot respond adequately to this variability.

Phenotypic Personalized Medicine (PPM), on the other hand, can guide mechanism-independent, patient-specific optimization of drug combinations. Based on experimental evidence, we established an individualized Phenotypic Response Surface (PRS) that provides a quantitative relationship, the PRS function, between the phenotypic outcome [e.g. tacrolimus serum trough level (TTL) in this study] and the drug dose.[14] Thus far, PRS-based PPM optimization has been demonstrated in *in vitro* and *in vivo* models of more than twenty different diseases, including the PPM clinical trials of organ transplantation, cancer, and infection. [25-29]

In a first-in-human clinical trial, we conducted a prospective pilot study with 4 PPM-dosed and 4 standard-of-care (SOC) dosed patients.[14]

PPM dynamically predicts the personalized tacrolimus dosing regimen over a time course. This is critical in the immediate post-transplant period when frequent changes to patient drug treatment regimens are common.

Comparisons between TTLs over the course of treatment showed that PPM dosing markedly improved the management of patient drug levels compared to SOC patients, including area-under-the-curve analyses, the frequency and magnitude of deviation from the target range, and the number of post-transplant days in the hospital. Large deviations (> 2.0 ng/mL) from trough target ranges were thought to be most important as they were indicators of possible adverse events (immunologic reactions for levels far below range and neuro- or nephrotoxicity for levels far above range).[14] Given these preliminary findings, we sought to conduct a larger prospective clinical trial to compare PPM dosing to SOC (clinician-dosing) in maintaining post-liver transplant patients within their clinically indicated target range.

## Methods

### Ethics/Regulatory Approval

The protocol was approved by the University of Florida Institutional Review Board (IRB201800053).

### Trial Design and Patient Population

In this single-center, pragmatic, randomized, partially blinded trial, adult participants who underwent primary or redo liver or simultaneous liver/kidney transplantation at the University of Florida Health were assigned immediately prior to the transplant procedure 1:1 to daily standard-of-care physician-guided dosing or PPM-guided dosing of tacrolimus. The study period covered from the day of liver transplantation to the day of discharge. No intervention was performed after discharge.

Inclusion criteria: 1) Subject undergoing primary or redo liver or simultaneous liver/kidney transplantation; 2) Subject or surrogate able to provide informed consent; 3) Subject at least 18 years of age.

Exclusion criteria: 1) Enrollment in another investigational device or drug study; 2) Subjects compulsorily detained; 3) Psychiatric or medical illness that may put subject at significant risk, confound study results, or interfere significantly with subject participation in study; 4) Patients with contraindications to tacrolimus or pre-operatively anticipated to be switched off tacrolimus.

### Randomization

To ensure equal sample sizes, subjects were assigned into either the PPM group or the SOC group using a permuted block randomization with a block size of 4. Subjects were blinded to group assignments; the clinical team was not blinded. The clinical team was blinded to block size. The clinical team screened and attempted to enroll all eligible patients undergoing liver transplantation. The principal investigator generated the randomization sequence and group assignment was assigned prior to the transplant operation to minimize bias from the operative course. To minimize dosing bias, the principal investigator was not involved in dosing of SOC subjects.

### Data Processing

In this study, the PPM dosing was initiated for the fourth day of tacrolimus administration, which is earlier than in the previous pilot study. [14] Due to the shorter timeframe, there were larger fluctuations in patient physiology, including rapid changes in liver and kidney function. Additionally, the number of dose changes in immunosuppression and other drugs required a fast decision on the PPM tacrolimus dosing. We have found that the next-day metabolism of tacrolimus is very close to that of the current day: i.e., TTL(t+1) / c(t+1) = TTL(t) / c(t)

Hence, the tacrolimus dose for next day, c(t+1). Is

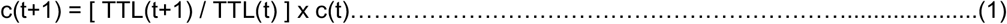

This means that for dynamically optimized dosing of tacrolimus, only the individual’s TTL and the tacrolimus dose from the previous day were needed to predict the tacrolimus dose for the next day. The tacrolimus doses of PPM patients were therefore modulated according to Function 1 to achieve the desired TTL within the clinically pre-designated target range.

### Clinical Protocol and Immunosuppression Summary

Following liver transplantation at the University of Florida, all study patients were started on SOC medication regimen per established center protocol: a three-drug combination of oral tacrolimus (Prograf), corticosteroid (intravenous methylprednisolone 500 mg iv intraoperatively followed by intravenous methylprednisolone 250 mg iv on post-operative days 1 and 2, 125 mg on post-operative day 3 and oral prednisone taper starting on post-operative day 4 if able to tolerate oral medications), and mycophenolate [usually oral mycophenolate mofetil (CellCept) 500 mg q12h or oral mycophenolic acid (Myfortic) 360 mg q 12h]. Induction was with intravenous methylprednisolone. Patients undergoing liver-kidney transplantation and those deemed at high risk for calcineurin-dependent renal injury also received basiliximab for induction and on post-operative day 4, starting tacrolimus on post-operative day 5-7 per clinical team discretion, and a lower tacrolimus target trough range. Blood TTLs were measured daily in the morning until patient discharge, using an automated chemiluminescent assay (CMIA; Abbott Laboratories) on the ARCHITECT i2000 platform. For the first 3 days that the patient was given tacrolimus, tacrolimus was dosed per SOC (usually capsule starting at 0.05 mg/kg administered orally or sublingually every twelve hours on day of transplantation) to allow enough data points to be gathered to allow PPM prediction. For SOC subjects, a transplant-specialized clinical pharmacist, together with the transplant-surgeon-on-call, continued to determine the subsequent doses. For PPM subjects, the daily treatment regimen details including drugs already administered, drugs to be administered, and hemodialysis or any other procedures to be performed were used for PPM analysis to provide the dosing from day 4. These data were sent to the PPM (prediction) team at UCLA. Following analysis, suggested tacrolimus doses were reviewed by a clinician and upon approval administered. The PPM process is not automated; PPM-suggested doses were based on safety limits set by the clinical team so that doses would always be within clinically-relevant levels. Patients remained on the trial until discharge from hospital at which point they reverted to SOC clinician-determined dosing. The standard target trough range was 8-10 ng/mL for the first month after transplant. This 2 ng/mL range was adjusted based on clinical condition of the patient per clinical team and institutional SOC. Reasons to adjust the trough range included risk/concern/evidence for infection, rejection, malignancy, or neurological or renal dysfunction. The decision to discharge the subjects were made by the clinical team based on adequate pain control, mobility and nutritional needs, therapeutic education, ongoing medical/surgical complications, and normal or normalizing liver function tests.[30]

### Primary and Secondary Endpoints

The primary endpoint measure was percent days with large (>2 ng/mL) deviation from target range during the initial post-transplant hospital stay, as this was deemed to be the closest correlate of dose-related adverse events. This was measured as the fraction of days dosed by either SOC or PPM where the resulting TTL the next day was >2 ng/mL outside the target range. The first 3 days where tacrolimus in both groups was dosed according to SOC were not included in the analysis. Most of this time, TTL was below the measurable range (< 3 ng/mL) and there were large variations of liver and kidney function.

The secondary endpoints included percent days outside-of-target range and mean area-under-the-curve outside-of-target-range per day.

Safety endpoints included biopsy-proven graft rejection, graft failure, death, infections, nephrotoxicity (biopsy proven acute kidney injury or calcineurin toxicity, anuria or oliguria requiring dialysis), or neurotoxicity (documented seizures, clinically significant tremors, or imaging-confirmed posterior reversible encephalopathy syndrome).

Exploratory/post hoc endpoints included LOS (length of stay), number of days for AST to normalize (DAST), and mean tacrolimus trough level during study period.

### Sample Size and Power Calculations

Based on previous results from the pilot study,[14] a sample size of 30 per group achieves 92% power to reject the null hypothesis of equal means when the population mean difference is 20% (31% vs. 11%) with standard deviations of 30% for SOC and 10% for PPM, and with a significance level of 0.05 using a two-sided two-sample unequal-variance t-test.

### Statistical Analyses

Demographic and clinical characteristics were summarized by treatment group using descriptive statistics, including mean (standard deviation) and median (first – third quartile, i.e., Q_1_-Q_3_). All subjects who received the allocated intervention (tacrolimus dosing using PPM versus SOC starting from day 4 of tacrolimus administration) were compared. Comparisons between the two groups were performed using either two-tailed Welch’s t-test or Student’s t-test. For non-normally distributed data, a two-tailed Wilcoxon Rank Sum test was used to compare medians. Receiver-operating-characteristic (ROC) analysis was used to find the optimal threshold to dichotomize length of stay after transplant and percent days out of target range for comparison. Spearman’s correlation was used to investigate the association between LOS and DAST. Statistical analyses were performed on JMP Pro 16.1.0 and confirmed with R 3.5.1. A Data and Safety Monitoring Board reviewed the study six months after first patient enrollment and every six months thereafter. The study was to be stopped in the event of a significant unacceptable difference between the two groups, as determined by the Board.

### Patient and Public Involvement

It was not appropriate or possible to involve patients or the public in the design, or conduct, or reporting, or dissemination plans of our research

## Results

Between September 1, 2018, and June 4, 2020, all adults scheduled to undergo primary, or redo liver or simultaneous liver/kidney transplantation were screened for eligibility. (Figure 1) 62 subjects met criteria and were randomized; 31 were assigned SOC-dosing and 31 were assigned PPM-dosing. Two patients in the PPM arm did not receive the intervention as their transplant operation was not performed after they were randomized and went to the operating room. Tacrolimus was discontinued for one patient in each arm due to neurotoxicity within the first few days after transplant, but as their tacrolimus levels were below target range, neurotoxicity was deemed not dose-related. Brain imaging revealed no evidence of tacrolimus neurotoxicity. Both patients were switched to cyclosporine, and thus there were no evaluable TTLs within the study period. One patient assigned to PPM dosing did not have tacrolimus level measured on post-operative day 5 due to a phlebotomy/laboratory error. As this would affect dosing accuracy and the study outcome measures, this patient was dropped from the study. One patient assigned to SOC refused multiple tacrolimus doses in the early post-operative period and was therefore dropped from the study. The remaining 56 (29 SOC and 27 PPM) patients underwent whole liver deceased unrelated-donor transplantation, completed the study, and were discharged from the hospital after transplantation. A total of 347 dosing decisions were made according to SOC in the control arm; 223 dosing recommendations were made by PPM. No PPM recommendations were overridden by the clinical team. The study was ended when the planned number of subjects were enrolled and completed the study.

**Figure 1.**
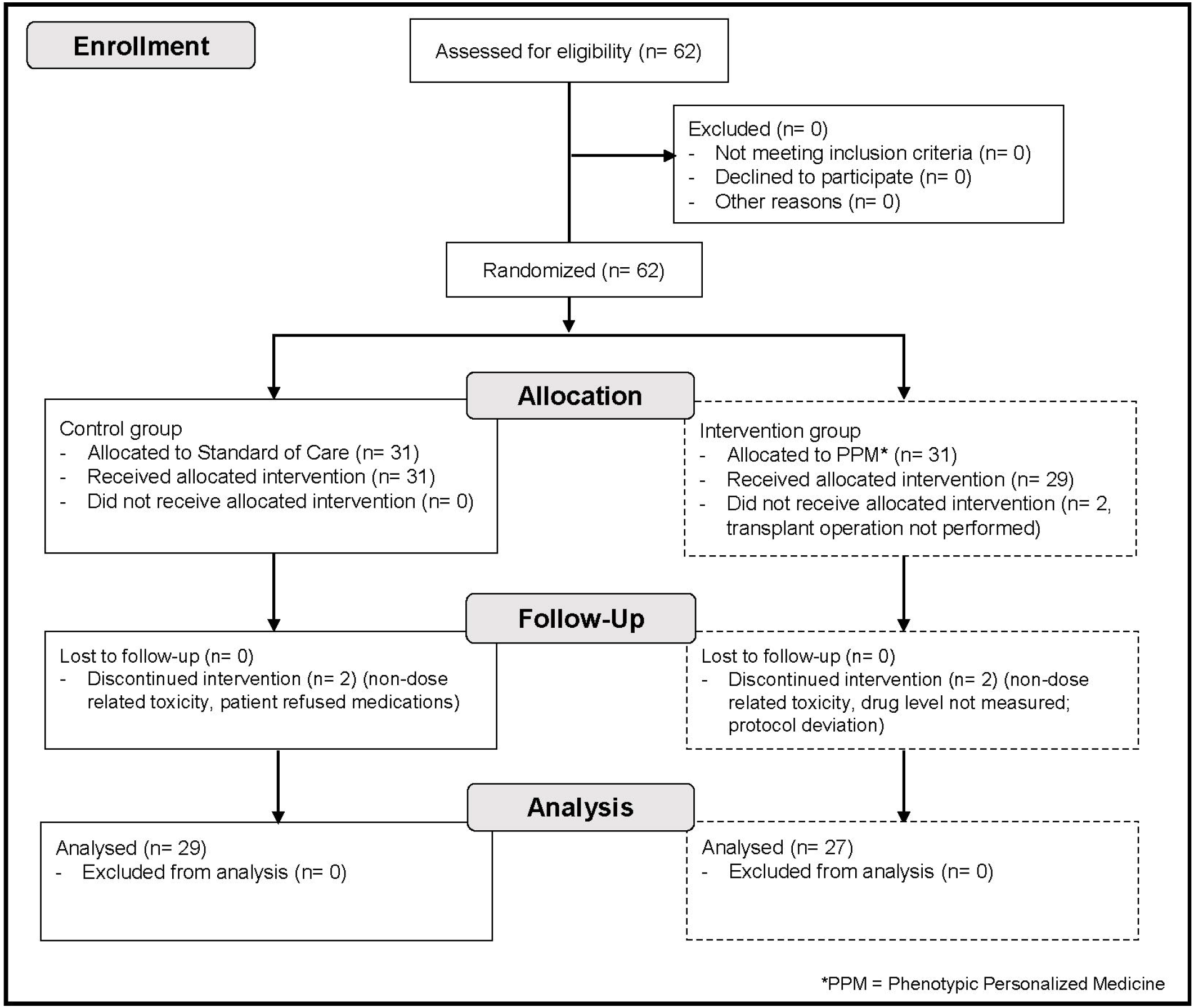
CONSORT Flow Diagram

In the study population of 56 patients, the median age was 58 (48-62); 32 patients (57%) were male. The median body-mass index (BMI) was 28 kg/m^2^ (25.1-32.1). The etiology for liver disease was alcohol-related in 21 patients (37.5%), non-alcoholic steatohepatitis in 13 patients (23%), and hepatitis C in 10 patients (18%). Ten patients (18%) had hepatocellular carcinoma, 2 (4%) underwent a redo liver transplantation, and 8 (14%) underwent simultaneous liver/kidney transplantation. No significant differences arose by chance in the baseline characteristics of the two groups. (Tables 1 & S1)

**Table 1.** Baseline characteristics of the analyzed study population HCC: Hepatocellular Carcinoma; SLKT: Simultaneous Liver-Kidney Transplant; OLT: Orthotropic Liver Transplant; BMI: Body Mass Index; NaMELD: Sodium-Model for End-stage Liver Disease, DCD: Donation after Circulatory Death

|  | Standard of Care<br>(n=29) | PPM<br>(n=27) |
| --- | --- | --- |
| Male | 16 (55%) | 16 (59%) |
| Female | 13 (45%) | 11 (41%) |
| HCC | 3 (10%) | 7 (26%) |
| SLKT | 5 (17%) | 3 (11%) |
| Redo OLT | 1 (3%) | 1 (4%) |
| Recipient Race/Ethnicity |  |  |
| Non-Hispanic White | 26 (90%) | 23 (85%) |
| Hispanic White | 1 (3%) | 4 (15%) |
| Non-Hispanic Black | 2 (6%) | 0 |
| Recipient Age (years) | 57 (41-61) | 58 (51-64) |
| BMI (kg/m<sup>2</sup>) | 27.9 (25.4-37.5) | 28.6 (23.3-32.6) |
| NaMELD | 27 (18.5-30.5) | 27 (15-32) |
| Warm Ischemia Time (min) | 31 (26.5-38.5) | 33 (26-39) |
| Cold Ischemia Time (min) | 390 (310-440) | 330 (300-420) |
| DCD Donor | 0 (0%) | 1 (4%) |
| Donor Age (years) | 41 (25.5-58) | 38 (32-54) |
| Hepatitis C Positive Donor | 1 (3%) | 2 (7%) |
| Donor Risk Index | 1.38 (1.25-2.01) | 1.35 (1.07-1.66) |
| Dialysis After Transplant | 3 (10%) | 2 (7%) |
| Donor Cause of Death |  |  |
| Anoxia | 8 (28%) | 11 (41%) |
| Cerebrovascular Accident | 11 (38%) | 10 (37%) |
| Trauma | 10 (34%) | 6 (22.2%) |
| Fluconazole Use After Transplant | 17 (59%) | 16 (59%) |
| Mycophenolic Acid Use | 2 (7%) | 2 (7%) |
| Basiliximab Use | 10 (34%) | 7 (25.9%) |
| Tacrolimus Target Range Other Than 8-10 ng/mL | 4 (14%) | 5 (19%) |

### Primary Endpoint

The primary endpoint was percent of study dosed days during the initial post-transplant hospitalization with TTL deviating by more than 2 ng/ml from the target range. The results showed a significant difference between the two groups. The mean percentage of post-transplant days with large deviations was 38.4 (27.4)% in the SOC group and 24.2 (19.1)% in the PPM group; (difference -14.1%, 95% CI: -26.7 to -1.5 %, P=0.029) (Figure 2A). The difference remains statistically significant with an intention-to-treat analysis: 41.4 (29.2)% in the SOC group and 24.2 (18.8)% in the PPM group (difference -17.2%, 95% CI: -29.7 to -4.6 %, P=0.0082).

**Figure 2.**
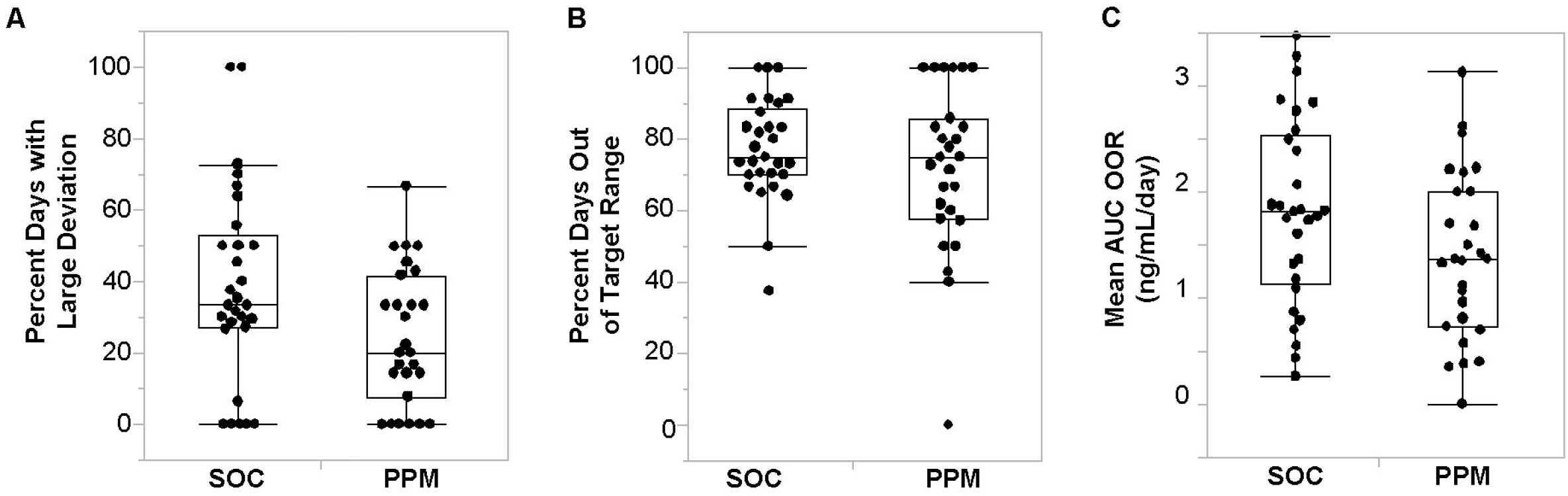
**(A)** Comparison of percent of dosed days with large deviations (>2 ng/ml) from target range during the post-transplant hospitalization between standard-of-care (SOC) and Phenotypic Personalized Medicine (PPM) dosing arms. PPM dosing resulted in a lower percent post-transplant days with *large deviations* from the target range compared to SOC dosing **(P=0.029). (B)** There were no statistically significant differences between the groups in the percent days with tacrolimus trough level outside-of-target-range (P=0.30). **(C)** There vvere also no statistically significant differences between the groups in the mean area-under-the-curve (AUC) outside-of-target-range (OOR) (P=0.087).

### Secondary Endpoints

One secondary endpoint was percent of study dosed days with deviations from the target range. Patients in the SOC group had a mean of 77.3 (14.2)% of post-transplant days with deviations from target range; the PPM group had 71.7 (23.2)% of post-transplant days with deviations; (difference -5.5%, 95% CI: -15% to 5.0%, P=0.30) (Figure 2B).

Another secondary endpoint was to quantify the degree to which the TTLs of each patient deviated from the target range by calculating the average the area-under-the-curve (AUC) outside-of-target-range (OOR) per day (Equation 3).

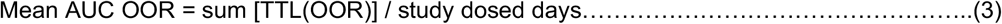

The difference in the mean AUC values between the study arms did not reach statistical significance (Figure 2C). SOC tacrolimus dosing led to a mean AUC OOR of 1.81 (0.88) ng/mL/day; PPM dosing had a mean of 1.40 (0.79) ng/mL/day (difference -0.41, 95% CI: -0.85 to 0.03, P=0.087).

### Safety Endpoints

The incidence of biopsy proven graft rejection episodes during the post-transplant hospitalization were similar (6 in each group; P=0.89). There were no graft failures or patient deaths within the one-year follow up of these patients.

### Exploratory Endpoints

Post-hoc analysis revealed that subjects in the SOC group had a 50% longer median length-of-stay (LOS) compared to the PPM group. The SOC group had a median LOS of 15 (10.5-20.5) days, while the PPM group had a median LOS of 10 (8-12) days (difference -5, 95% CI: -2 to -8, P=0.0026) (Figure 3).

**Figure 3.**
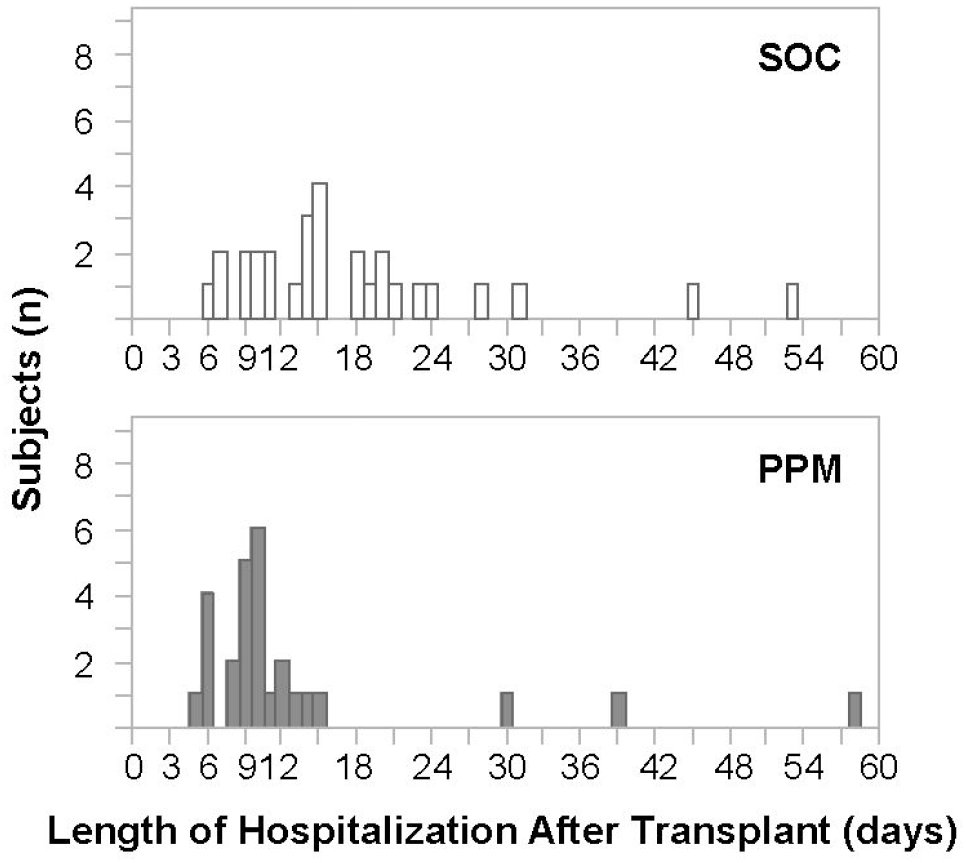
Patients in the SOC group had a longer median length of stay (LOS) after transplantation of 15 (10.5-20.5) days; the PPM group had a median LOS of 10 (8-12) days **(P=0.0026)**.

The mean TTL of the SOC group was lower than the PPM group (and within the commonly used target range of 8-10 ng/mL) [7.6 (1.6) ng/mL versus 8.4 (1.4), difference 0.8, 95% CI: 0.002 to 1.6, P=0.034]. (Figure 4A) This difference in the mean TTLs prompted us to explore systematic differences between the two dosing methods.

**Figure 4.**
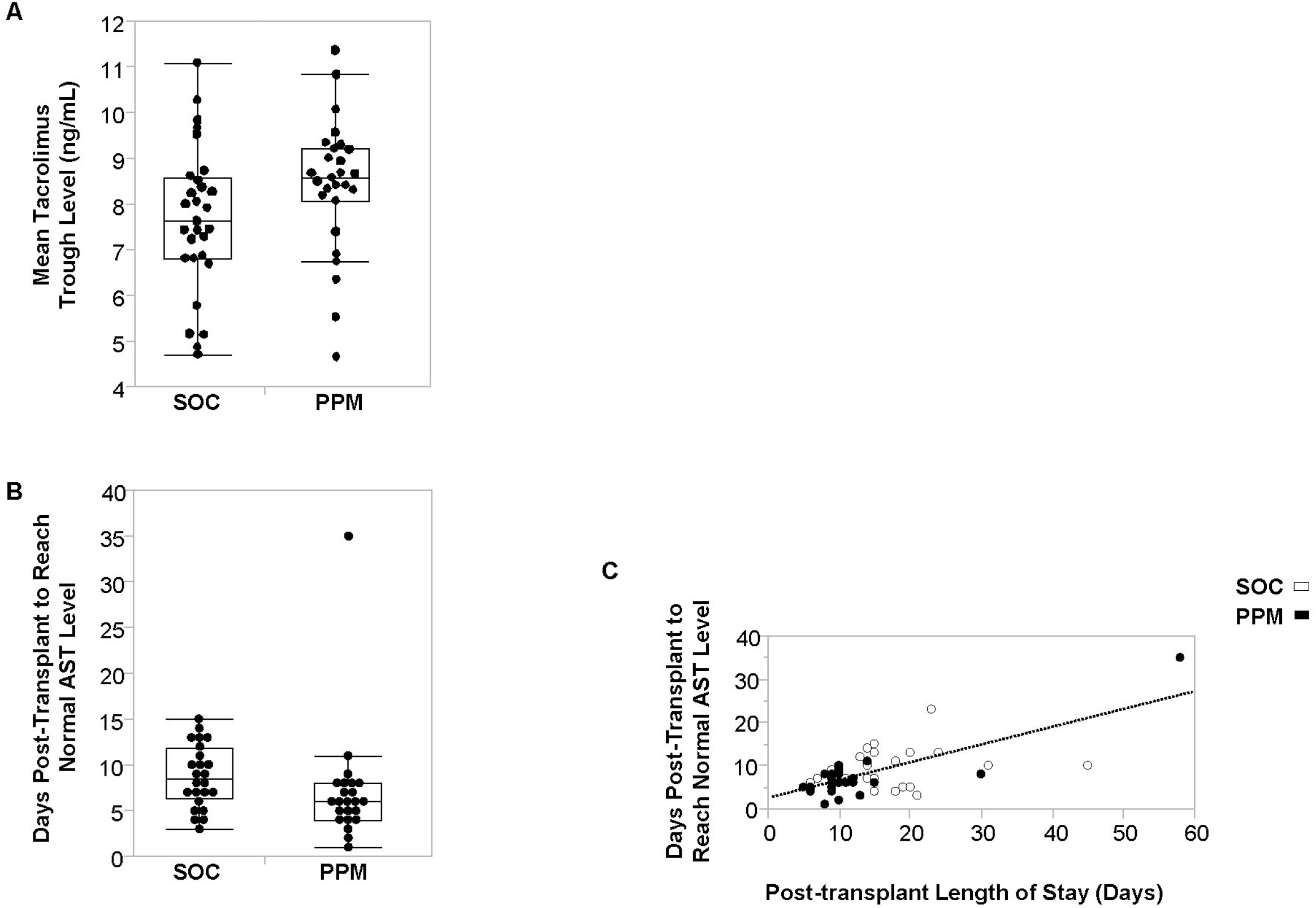
**(A)** The mean tacrolimus trough level in the SOC group was 7.6 (1.6) ng/ml; in the PPM group it was 8.4 (1.5) ng/ml **(P=0.034). (B)** Patients in the SOC group took a median of 8.5 days (Q_1_-Q_3_ 6.25-11.75) to normalize aspartate aminotransferase (AST) levels. The PPM group reached normal AST levels at 6 days (Q_1_-Q_3_4-8) **(P=0.014). (C)** Length of stay after transplantation was directly correlated with the number of days it took for AST levels to normalize (R^2^=0.42, **P<0.0001)**.

We therefore tested whether either method was consistently under- or over-dosing patients. With regard to underdosing (i.e., TTLs below target range), the mean AUC below target range (i.e., 8 ng/mL minus TTL) is a measure of the chance of suffering graft injury, consistent with previous findings correlating tacrolimus concentration and liver graft integrity markers.[12] We found that the SOC group had a statistically significantly larger mean AUC below target range than the PPM group [1.4 (1.0) ng/mL/day versus 0.76 (0.51) ng/mL/day, respectively (difference -0.64, 95% CI: -1.05 to -0.23, P=0.0096)] (Figure S1A).

As for overdosing, the mean AUC above target range (i.e., TTL minus 10 ng/mL) is a measure of the chance of having drug toxicity. There was no significant difference in mean AUC above target range between the two groups [SOC: 0.42 (0.47) ng/mL/day versus PPM: 0.64 (0.58) ng/mL/day, (difference 0.22, 95% CI: -0.07 to 0.50, P=0.13)] (Figure S1B).

To investigate the relationship between recovery from transplant-related liver injury and LOS, we analyzed the number of days it took for AST levels (a standard measure of liver injury) to normalize (DAST). The PPM group reached normal AST levels more quickly than the SOC group [PPM: 6 days (4-8); SOC: 8.5 days (6.25-11.75)]; P=0.014 (Figure 4B). Six patients, three in each group, whose AST levels did not reach normal levels prior to discharge, were excluded from this analysis. While there was a linear relationship between DAST and LOS (R^2^=0.42, P<0.0001) (Figure 4C), post liver transplant patient discharge is mainly based on qualitative phenotypes, not DAST. Hence, the scatters are fairly large.

To control for the impact of LOS on measures of tacrolimus dosing, we reanalyzed the data using a standard timeframe of only up to ten days after transplantation (i.e., first three days of lead-in standard-of-care dosing and then seven days of study dosing). This time period was chosen to optimize the inclusion of patients with short hospital stays (more than 75% of the patients had a LOS of 10 days or longer) while minimizing the influence of long-term stabilization of tacrolimus dosing. During this period, patients in the SOC group had a mean of 42.1 (30.9)% of post-transplant days with large deviations from target range; the PPM group had 24.9 (19.5)% of post-transplant days with large deviations; (difference -17.3%, 95% CI: -3.5% to -31%, P=0.015). There was no statistically significant difference during that timeframe for percent post-transplant days with deviations from target range (SOC 78.6% versus PPM 72.9%, difference -5.7%, 95% CI: -17.7% to 6.3%, P=0.35). However, there was a significant difference for the AUC-OOR per day within this timeframe (SOC 2.01 ng/mL/day versus PPM 1.41 ng/mL/day, difference -0.60, 95% CI: -1.15 to -0.04, P=0.035)

## Discussion

Tacrolimus dosing in the early period after liver transplantation is a challenging task that involves factors other than dose alone; it includes the types and doses of other medications, liver and kidney function, metabolism, pharmacogenetics, and other factors. The important task of getting tacrolimus blood levels to the therapeutic range without under- or overdosing is managed daily by the clinical team. This team typically includes transplant pharmacists and physicians who together incorporate a combination of factors such as organ function, recovery of bowel function and absorption, and concomitant medications. This complex system is difficult to navigate and has not lent itself well to predictive protocols or algorithms.

The PPM platform functions entirely using quantitative phenotypic data (tacrolimus trough level in this current example) to personalize treatment. As a result, it is applicable and valuable in systems of varying complexity, such as in this trial where PPM navigated the complex biological, physiological, and clinical context of post-transplant tacrolimus dosing. The discovery of preservation of metabolism ratio from one day to the next, Function-1, represents a directly actionable approach towards improved immunosuppression therapy.

In this study, we found that PPM dosing outperformed standard-of-care (SOC) clinician-determined dosing by decreasing the percent of days with large deviations from the target blood trough level range in daily dosing of tacrolimus after liver and liver/kidney transplantation. While previous studies have explored predictive diagnostic and prognostic technologies, no prospective trials have assessed the ability to affect and measurably improve the routine clinical care of patients.[31, 32] This study represents the first such in-human application in a randomized clinical trial. It confirmed the findings of the pilot study with a larger, independent, and more heterogeneous cohort, at a different institution and with statistically significant differences.[14]

The primary endpoint of this study was the percent of days with a large deviation from the target trough drug level range, which was decreased in the PPM group compared to the SOC group (Figure 2A). PPM also improved short-term clinical outcomes and presumably reduced cost in that patients in the PPM dosing arm were discharged five days earlier that the SOC arm (Figure 3), a finding that replicated the results of the pilot study.[14]

One potential explanation of how better dosing led to shorter LOS is hinted at by the additional finding that PPM patients had a faster time to normalization of AST (Figure 4B). Oellerich et al. have previously reported that tacrolimus trough concentrations in the short-term after liver transplantation are associated with graft integrity. It therefore follows that the higher in-range TTL of the PPM patients (Figure 4B) can lead to improved post-transplant liver integrity, as measure by a more rapid normalization of AST. Not only is TTL higher in PPM-dosed patients overall, but SOC patients are underdosed more than PPM-dosed patients (Figure S1A).

Preserving stable tacrolimus trough levels early after solid organ transplantation is a complex, yet crucial, task, as reducing the magnitude of trough level variability and the period of time in which large variations occur has a significant impact on post-transplant outcomes.[12, 33] Multiple studies have found mean tacrolimus trough concentrations within therapeutic range during the first 15 days after liver transplantation to be associated with lower risk of graft loss when compared to under- and overdosing.[13] This was suggested to be linked with the significantly higher incidence of neurologic, cardiovascular, and acute renal failure complications in the high variability group. Furthermore, high intra-patient tacrolimus level variability in the first 30 post-transplant days has been correlated with significantly worse one-year and long-term graft survival.[34] Therefore, the decrease in percent days with a large deviation from target levels is a clinically significant step towards improving graft and patient outcomes in liver transplantation.

Though this is a randomized prospective clinical trial and performed at an independent institution from the pilot trial[14], it is still limited by being a single-center study and its results will have to be replicated at multiple sites and larger populations. It was not powered to assess differences in clinical outcomes. In addition, given the post hoc nature of the LOS and DAST difference analysis, these findings will need further validation, especially as some subjects reached DAST before study dosing initiation. However, this study does provide evidence that systematic drug dosing using PPM can augment the clinical care of patients prospectively and can lead to daily recommendations that can respond dynamically to individual patient circumstances.

## Data Availability

Deidentified participant data, study protocol, statistical analysis plan, and informed consent form will be available with publication from AZ upon reasonable request and after approval of a proposal with a signed data access agreement.

## Abbreviations

AST: aspartate aminotransferase
c(t): tacrolimus dose on day t
DAST: day at which the AST level normalized
LOS: length of stay
OOR: outside of target range
PPM: Phenotypic Personalized Medicine
PRS: parabolic response surface
ROC: receiver operating characteristic
SOC: standard of care
TTL(t): tacrolimus trough level on day t

## Contributors

AZ and CMH designed the study. AZ, UBK, CW, SS, KAA, MWJ, NRB, SD, TB, and DMM participated in the conduct of the study, data acquisition, analysis, and interpretation. CMH, JK, ML, and AZ developed and performed the PPM analysis and made dose recommendations. JK, ML, JHL, DML, CMH, and AZ accessed and verified the data and did the statistical analysis. AZ, SD, and CMH wrote the manuscript. All authors critically reviewed the manuscript.

## Trial Registration

Optimizing Immunosuppression Drug Dosing Via Phenotypic Precision ClinicalTrials.gov (NCT03527238).

## Data sharing

Deidentified participant data, study protocol, statistical analysis plan, and informed consent form will be available with publication from AZ after approval of a proposal with a signed data access agreement.

