## Supplemental Table 1 for "Personalized Tacrolimus Dosing After Liver Transplantation: A Randomized Clinical Trial"

|  | Standard of Care | PPM |
| --- | --- | --- |
|  | (n=31) | (n=31) |
| Male | 17 (55%) | 18 (58%) |
| Female | 14 (45%) | 13 (42%) |
| HCC | 3 (10%) | 8 (26%) |
| SLKT | 6 (19%) | 3 (10%) |
| Redo OLT | 1 (3%) | 2 (6%) |
| Recipient Race/Ethnicity |  |  |
| Non-Hispanic White | 27 (87%) | 26 (84%) |
| Hispanic White | 2 (6%) | 4 (13%) |
| Non-Hispanic Black | 2 (6%) | 0 |
| Asian | 0 | 1 (3%) |
| Recipient Age | 57 (44.5-61) | 58 (50.5-63) |
| BMI (kg/m^2^) | 27.7 (25-31.1) | 28.6 (23-32.4) |
| NaMELD | 27 (19-29) | 27 (16-30.5) |

**Table S1.** Baseline characteristics of the randomized population

HCC: Hepatocellular Carcinoma; SLKT: Simultaneous Liver-Kidney Transplant; OLT: Orthotropic Liver Transplant; BMI: Body Mass Index; NaMELD: Sodium-Model for End-stage Liver Disease
