## Supplementary figures and images for "Personalized Tacrolimus Dosing After Liver Transplantation: A Randomized Clinical Trial"

### Supplemental Figure 1

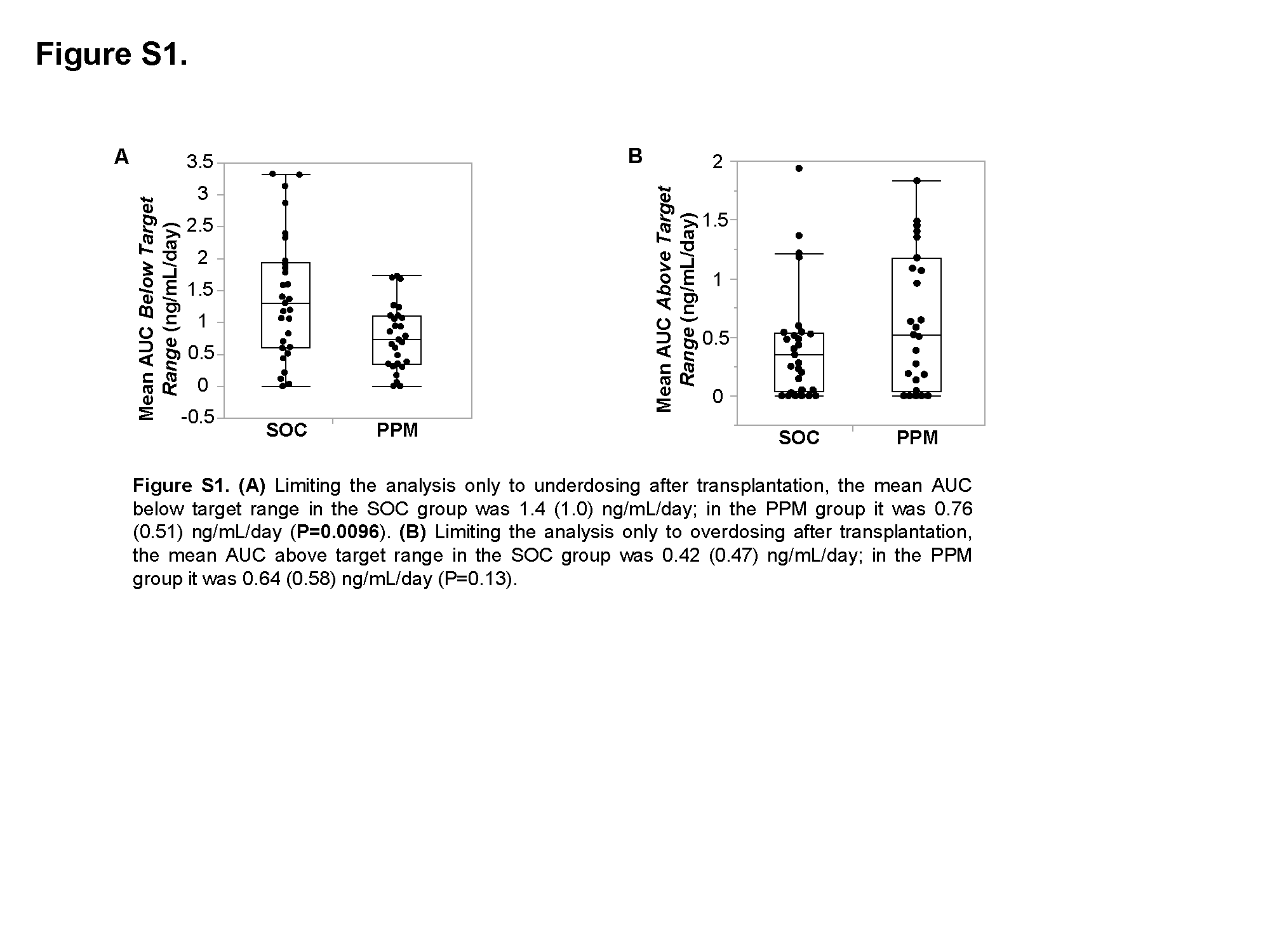
